# Excellent recanalization and small core volumes are associated with favorable AMPAC score in patients with acute ischemic stroke secondary to large vessel occlusion

**DOI:** 10.1101/2023.08.27.23294705

**Authors:** Vivek Yedavalli, Manisha Koneru, Meisam Hoseinyazdi, Karen Copeland, Risheng Xu, Licia Luna, Justin Caplan, Adam Dmytriw, Adrien Guenego, Jeremy Heit, Gregory Albers, Max Wintermark, Fernando Gonzalez, Victor Urrutia, Judy Huang, Richard Leigh, Elisabeth Marsh, Rafael Llinas, Marlis Gonzalez Hernandez, Argye Hillis

## Abstract

**Background and aim:** Acute ischemic stroke due to large vessel occlusion (AIS-LVO) is a major cause of functional dependence, an important determinant of discharge disposition. The aim of this study is to assess the utility of pretreatment and interventional parameters as predictors of favorable Activity Measure for Post Acute Care (AMPAC) scores for optimal discharge planning.

**Methods:** In this retrospective multicenter analysis, inclusion criteria were as follows: a) CT angiography (CTA) confirmed LVO from 9/1/2017 to 9/22/2022; b) diagnostic CT perfusion; and c) available AMPAC scores. Patients were then dichotomized into favorable and unfavorable AMPAC for analysis. A multivariate logistic regression was performed using specific variables that were clinically relevant and significant on univariate regression analyses. A receiver operator characteristics (ROC) analysis was then performed to assess the diagnostic performance of the logistic regression model. A p value of <= 0.05 was considered significant.

**Results:** In total, 229 patients (mean +-SD 70.65 +-15.2 [55.9% female]) met our inclusion criteria. Favorable AMPAC patients were younger (61.3 versus 70.7, p < 0.0001), had lower admission glucose (mean, 124.19 versus 136.83, p = 0.042), lower blood urea nitrogen (mean, 15.59 versus 19.11, p = 0.0009), and lower admission National Institutes of Health Stroke Scale (NIHSS) (mean, 10.58 versus 16.15, p < 0.0001). Multivariate regression analyses revealed age, admission NIHSS, relative cerebral blood flow (rCBF) < 30% volume, and modified thrombolysis in cerebral infarction (mTICI) score to be independent predictors of favorable AMPAC (p<0.047 for all predictors). ROC analysis of the combined model revealed an area under the curve (AUC) of 0.83 (IQR 0.75 - 0.86).

**Conclusion:** Excellent recanalization, smaller core volumes, younger age and lower stroke severity independently predict favorable outcomes as measured by AMPAC. Our study further emphasizes the significance of minimizing core volume and aiming for excellent recanalization in order to optimize discharge disposition in AIS-LVO patients.

## Introduction

Large vessel occlusions (LVOs) cause acute ischemic stroke (AIS) in up to 46% of patients^1^ and are a leading cause of morbidity across the world^2^. Patients presenting with AIS caused by LVO (AIS-LVO) have disproportionately increased functional limitations compared to non-LVO AIS patients^3^, underscoring the importance of timely treatment in these patients. Functional limitations, as a result of an AIS patient’s hospital stay, is an important determinant of discharge location^4,5^. Although functional limitations are an important factor, there are several other factors that affect the patient’s discharge disposition after acute hospital admission^4^. However, there is still no clinical consensus on which metrics are most reliable in determining the optimal discharge location in poststroke patients ^4,6^. The challenges with the discharge location decision making process emphasize the importance of investigating the utility of predictive biomarkers as additional supportive data points for consideration.

The Activity Measure for Post-Acute Care (AMPAC) score is a reliable and easy to perform validated metric that measures daily activity and basic mobility^7,8^. AMPAC scores are predictive of not only discharge location^4^ but 30-day hospital readmission^7^ and functional outcomes^9^ in poststroke care assessments. Furthermore, AMPAC scores are utilized to determine the most appropriate type of discharge facility^4^ and minimize hospital readmission when preventable^7^. For the aforementioned reasons, AMPAC is now increasingly used with AIS-LVO patients as a unique assessment of in-hospital activity for discharge status determination^10–12^.

For AIS-LVO patients, baseline neuroimaging with CT is an important component within the overall workup. Pre-treatment comprehensive CT imaging consisting of noncontrast CT (NCCT), CT angiography, and CT perfusion (CTP) provides information on the ischemic core, salvageable tissue or penumbra, and collateral status (CS) ^13^. The valuable information given by the CT evaluation aids in the decision-making process with administering reperfusion therapies - namely IV thrombolysis (IV tPA), mechanical thrombectomy (MT), or both. Prior landmark trials have validated the use of perfusion imaging in determining MT eligibility, demonstrating improved outcomes for up to 24 hours after symptom onset^14,15^. Nevertheless, the potential utility of pretreatment comprehensive CT imaging, in conjunction with clinical and demographic factors, in determining post-acute care discharge needs for AIS-LVO patients has not been explored to date.

The purpose of our study is to determine which pretreatment and interventional parameters are predictive of favorable AMPAC scores in patients presenting with AIS-LVO with a focus on pretreatment CT imaging. We hypothesize that patients with smaller baseline ischemic cores are associated with favorable AMPAC daily activity and basic mobility scores.

## Methods

### Population

We identified patients with confirmed anterior circulation LVOs on CTA using baseline comprehensive CT evaluation (which includes NCCT, CTA, CTP) from three centers within the Johns Hopkins Medical Enterprise (Johns Hopkins Hospital - East Baltimore, Bayview Medical Campus, and Suburban Hospital). The Johns Hopkins East Baltimore and Bayview campuses are accredited comprehensive stroke centers. This study was approved through the Johns Hopkins School of Medicine institutional review board (JHU-IRB00269637). In this retrospective multicenter analysis, inclusion criteria were as follows: a) CT angiography (CTA) confirmed LVO from 9/1/2017 to 9/22/2022; b) diagnostic CT perfusion; and c) available AMPAC scores.

### Data Collection

Baseline and clinical data collected for each patient included demographics, risk factors for AIS (including diabetes mellitus, hypertension, coronary artery disease, atrial fibrillation), admission glucose, admission NIH stroke scale (admission NIHSS), admission blood urea nitrogen (BUN), admission creatinine, admission hemoglobin (Hb), Alberta Stroke Program Early CT Score (ASPECTS) scores, site of occlusion, and laterality of occlusion. Additional collected parameters include number of passes, recanalization time, modified thrombolysis in cerebral infarction (mTICI) score; presence of complication such as hemorrhagic transformation (HT) of HI subtype only as defined by the European Cooperative Acute Stroke Study (ECASS) 2 trial^16^. ASPECTS scores were calculated and baseline CTAs were reviewed for presence and site of LVO by an experienced neuroradiologist (VSY, 6 years of experience). Treatment type including IV tPA, MT, or both were noted. Patients were then dichotomized into favorable (defined as a daily activity score >= 19 and basic mobility score of >= 17) ^7^ and unfavorable AMPAC (defined as a daily activity score < 19 and basic mobility score of < 17)^7^ for analysis. Patients who have favorable AMPAC scores in daily activity or basic mobility assessments, but not in both, were categorized as unfavorable.

#### Imaging Analysis

Whole brain pretreatment CTP was performed on the Siemens Somatom Force (Erlangen, Germany) with the following parameters: 70 kVP, 200 Effective mAs, Rotation Time 0.25 s, Average Acquisition Time 60 s, Collimation 48 × 1.2 mm, Pitch Value 0.7, 4D Range 114 mm x 1.5 seconds. CTP images are then post-processed using RAPID commercial software (IschemaView, Menlo Park) for generating quantitative relative cerebral blood flow (rCBF) and time to maximum (Tmax) volumes as well as qualitative Tmax maps. Hypoperfusion index ratio (HIR) was calculated as the ratio of the Tmax > 10 seconds and Tmax > 6 seconds volumes^17^. An HIR of 0.4 and below is deemed robust CS^18^.

#### Clinical Outcomes Assessment

AMPAC scores were determined by the certified physical and occupational therapists providing clinical care during admission.

#### Outcome Measures

The primary outcomes were favorable AMPAC, defined as a daily activity score >= 19 and basic mobility score of >= 17^7^.

### Statistical analysis

The collected data were coded, tabulated, and statistically analyzed using IBM SPSS statistics (Statistical Package for Social Sciences) software version 28.0 (IBM Corp., Chicago, USA, 2021). Quantitative data were tested for normality using Shapiro-Wilk test, described as mean and standard deviation (SD), and compared using the two-sided Student’s t-test. If data were not normally distributed, they were described as median with interquartile ranges (IQR), and compared using the Mann Whitney test. Categorical variables were reported as frequencies and compared using the likelihood ratio test. Univariate and multivariate regression analyses for predicting favorable AMPAC scores were performed. A multivariate logistic regression was built using statistically significant univariate predictors and pre-specified clinical factors. The multivariate model was refined with elimination of non-significant parameters with the lowest effect size, yielding a model with four clinical parameters and three imaging parameters. Receiver operating characteristics (ROC) curve with area under the curve (AUC) was used to evaluate model performance. A p-value ≤ 0.05 was significant.

## Results

In total, 229 patients (mean +-SD 70.65 +-15.2 [55.9% female]) met our inclusion criteria with 79 (79/229, 34.5%) in the favorable and 150 (150/229, 65.5%) in the unfavorable groups, respectively.

Favorable AMPAC score patients were younger (61.3 versus 70.7, p < 0.0001), had lower admission glucose (mean, 124.19 versus 136.83, p = 0.042), lower BUN (mean, 15.59 versus 19.11, p = 0.0009), and lower admission NIHSS (mean, 10.58 versus 16.15, p < 0.0001; Table 1).

**Table 1:** Baseline demographic and clinical characteristics for patients with either favorable or unfavorable AMPAC scores.

| Characteristics | All (n=229) | Favorable AMPAC Score (n=79) | Unfavorable AMPAC Score (n=150) | p-value |
| --- | --- | --- | --- | --- |
| *Demographic* |  |  |  |  |
| Age (years), mean (SD) | 67.43 (15.50) | 61.32 (14.42) | 70.65 (15.12) | **<.0001** |
| Sex, no. (%) |  |  |  |  |
| Female | 128 (55.90%) | 45 (56.96%) | 83 (55.33%) | 0.8134 |
| Male | 101 (44.10%) | 34 (43.04%) | 67 (44.67%) |  |
| Race, no. (%) |  |  |  |  |
| Black/African American | 97 (42.36%) | 31 (39.24%) | 66 (44.00%) | 0.7419 |
| Caucasian | 120 (52.40%) | 44 (55.70%) | 76 (50.67%) |  |
| Asian | 5 (2.18%) | 1 (1.27%) | 4 (2.67%) |  |
| Other | 7 (3.06%) | 3 (3.80%) | 4 (2.67%) |  |
| Tobacco Use, no. (%) | 100 (43.67%) | 34 (43.04%) | 66 (44.00%) | 0.8890 |
| Hypertension, no. (%) | 179 (78.17%) | 57 (72.15%) | 122 (81.33%) | 0.1134 |
| Dyslipidemia, no. (%) | 118 (51.53%) | 39 (49.37%) | 79 (52.67%) | 0.6349 |
| Diabetes Mellitus, no. (%) | 59 (25.76%) | 16 (20.25%) | 43 (28.67%) | 0.1608 |
| Heart Disease, no. (%) | 117 (51.09%) | 34 (43.04%) | 83 (55.33%) | 0.0765 |
| Atrial Fibrillation, no. (%) | 89 (38.86%) | 25 (31.65%) | 64 (42.67%) | 0.1014 |
| Prior Stroke/Transient Ischemic Attack, no. (%) | 42 (18.34%) | 14 (17.72%) | 28 (18.67%) | 0.8602 |
| BMI (kg/m<sup>2</sup>), mean (SD) | 29.38 (7.91) | 29.82 (7.70) | 29.15 (8.04) | 0.5438 |
| SBP (mmHg), mean (SD) | 152.28 (29.11) | 149.92 (28.51) | 153.52 (29.43) | 0.3710 |
| DBP (mmHg), mean (SD) | 87.62 (20.30) | 87.01 (20.02) | 87.93 (20.51) | 0.7443 |
| HR (bpm), mean (SD) | 85.10 (20.62) | 82.78 (16.07) | 86.31 (22.61) | 0.1736 |
| Glucose (mg/dL), mean (SD) | 132.47 (52.54) | 124.19 (33.59) | 136.83 (59.82) | **0.0418** |
| BUN (mg/dL), mean (SD) | 17.90 (9.02) | 15.59 (5.69) | 19.11 (10.17) | **0.0009** |
| Cr (mg/dL), mean (SD) | 1.09 (0.61) | 1.01 (0.32) | 1.13 (0.72) | 0.0812 |
| Hemoglobin (g/L), mean (SD) | 12.97 (1.94) | 12.80 (1.85) | 13.05 (1.99) | 0.3462 |
| *Stroke Characteristics* |  |  |  |  |
| Admission NIHSS, mean (SD) | 14.22 (6.86) | 10.58 (6.61) | 16.15 (6.20) | **<.0001** |
| Premorbid mRS, no. (%) | n=228 | n=79 | n=150 |  |
| 0 | 87 (38.16%) | 49 (67.12%) | 87 (58.00%) | 0.1483 |
| 1 | 19 (8.33%) | 14 (19.18%) | 19 (12.67%) |  |
| 2 | 7 (3.07%) | 4 (5.48%) | 7 (4.67%) |  |
| 3 | 24 (10.53%) | 4 (5.48%) | 24 (16.00%) |  |
| 4 | 0 (0.00%) | 0 (0.00%) | 0 (0.00%) |  |
| 5 | 0 (0.00%) | 0 (0.00%) | 0 (0.00%) |  |
| Stroke Etiology (TOAST Criteria), no. (%) |  |  |  |  |
| Larger Artery Atherosclerosis | 39 (17.18%) | 12 (15.38%) | 27 (18.12%) | 0.2693 |
| Cardioembolism | 117 (51.54%) | 35 (44.87%) | 82 (55.03%) |  |
| Small-Vessel Occlusion | 0 (0.00%) | 0 (0.00%) | 0 (0.00%) |  |
| Stroke of Other Determined Etiology | 12 (5.29%) | 5 (6.41%) | 7 (4.70%) |  |
| Stroke of Undetermined Etiology | 59 (25.99%) | 26 (33.33%) | 33 (22.15%) |  |
| Laterality, no. (%) |  |  |  |  |
| Left | 108 (47.16%) | 35 (44.30%) | 73 (48.67%) | 0.2962 |
| Right | 120 (52.40%) | 43 (54.43%) | 77 (51.33%) |  |
| Bilateral | 1 (0.44%) | 1 (1.27%) | 0 (0.00%) |  |
| Occlusion Site, no. (%) |  |  |  |  |
| ICA | 8 (2.64%) | 2 (2.60%) | 6 (2.67%) | 0.9943 |
| M1 | 163 (71.81%) | 55 (71.43%) | 108 (72.00%) |  |
| M2 | 58 (25.55%) | 20 (25.97%) | 38 (25.33%) |  |
| IV tPA, no. (%) | 62 (27.07%) | 24 (30.38%) | 38 (25.33%) | 0.4165 |
| MT Not Attempted, no. (%) | 48 (20.96%) | 20 (25.32%) | 28 (18.67%) | 0.0773 |
| mTICI, no. (%) | n=181 | n=59 | n=122 |  |
| 0 | 8 (4.42%) | 1 (1.69%) | 7 (5.74%) |  |
| 1 | 4 (2.21%) | 1 (1.69%) | 3 (2.46%) |  |
| 2A | 7 (3.87%) | 1 (1.69%) | 6 (4.92%) |  |
| 2B | 42 (23.20%) | 9 (15.25%) | 33 (27.05%) |  |
| 2C | 28 (15.47%) | 8 (13.56%) | 20 (16.39%) |  |
| 3 | 92 (50.83%) | 39 (66.10%) | 53 (43.44%) |  |

On pretreatment imaging, patients with favorable AMPAC patients had significantly lower rCBF and Tmax volumes (p<0.049 for all parameters; Table 2).

**Table 2:** Imaging characteristics for patients with either favorable or unfavorable AMPAC scores.

| Characteristics | All (n=229) | Favorable AMPAC Score (n=79) | Unfavorable AMPAC Score (n=150) | p-value |
| --- | --- | --- | --- | --- |
| Calculated Tmax > 4s volume (mL), median (IQR) | 214 (139.75-323.25) | 190 (123-247) | 214 (139.75-323.25) | **0.0406** |
| Calculated Tmax > 6s volume (mL), median (IQR) | 117 (65.25-164.75) | 97 (51.5-141) | 117 (65.25-164.75) | **0.0343** |
| Calculated Tmax > 8s<br>volume (mL), median (IQR) | 69.5 (36-116.75) | 54 (20.5-96) | 69.5 (36-116.75) | **0.0279** |
| Calculated Tmax > 10s<br>volume (mL), median (IQR) | 40 (13.5-95.75) | 34 (6.5,-69.5) | 40 (13.5-95.75) | **0.0257** |
| Calculated rCBF < 20%<br>(mL), median (IQR) | 0 (0-13.75) | 0 (0-6) | 0 (0-13.75) | **0.0205** |
| Calculated rCBF < 30%<br>(mL), median (IQR) | 8.5 (0-41) | 0 (0-16.5) | 8.5 (0-41) | **0.0010** |
| Calculated rCBF < 34%<br>(mL), median (IQR) | 14.5 (0-53.75) | 0 (0-23.5) | 14.5 (0-53.75) | **<.0001** |
| Calculated rCBF < 38%<br>(mL), median (IQR) | 22.5 (5-62) | 5 (0-29.5) | 22.5 (5-62) | **0.0004** |
| Hypoperfusion Intensity<br>Ratio, median (IQR) | 0.4 (0.2-0.55) | 0.3 (0.15-0.5) | 0.4 (0.2-0.6) | **0.0449** |
| CBV Index, median (IQR) | 0.8 (0.7-0.9) | 0.8 (0.7-1.0) | 0.8 (0.7-0.9) | **0.0044** |

Multivariate logistic regression analyses revealed age (p < 0.0001), admission NIHSS (p < 0.0001), mTICI score (p = 0.0382), and rCBF < 30% volume (p = 0.0465), to be independent predictors of favorable AMPAC (Table 3). Admission glucose (p = 0.062) and female sex (p = 0.310) approached significance in favorable AMPAC prediction. The ROC curve for the combined model revealed an AUC of 0.83 (95% confidence interval 0.75 - 0.86; Figure 1).

**Table 3:** Odds ratio for predictors included in multivariate logistic regression model for predicting favorable over unfavorable AMPAC scores.

| **Characteristic** | **Odds Ratio (95% CI)** | **p-value** |
| --- | --- | --- |
| Age (per year) | 0.94 (0.92-0.96) | **<0.0001** |
| Admission NIHSS (per unit) | 0.88 (0.83-0.93) | **<0.0001** |
| Admission Glucose (per mg/dL) | 0.99 (0.98-1.00) | 0.0618 |
| Sex |  |  |
| Male | Reference | 0.3103 |
| Female | 1.43 (0.72-2.83) |  |
| rCBF <30% Volume (per mL) | 0.99 (0.97-0.99) | **0.0465** |
| mTICI |  |  |
| 0/1/2A/2B | Reference | **0.0382** |
| 2C/3 | 3.06 (1.24-7.52) |  |

**AUC = 0.83 95%CI: 0.75 – 0.86**

**Figure 1:**
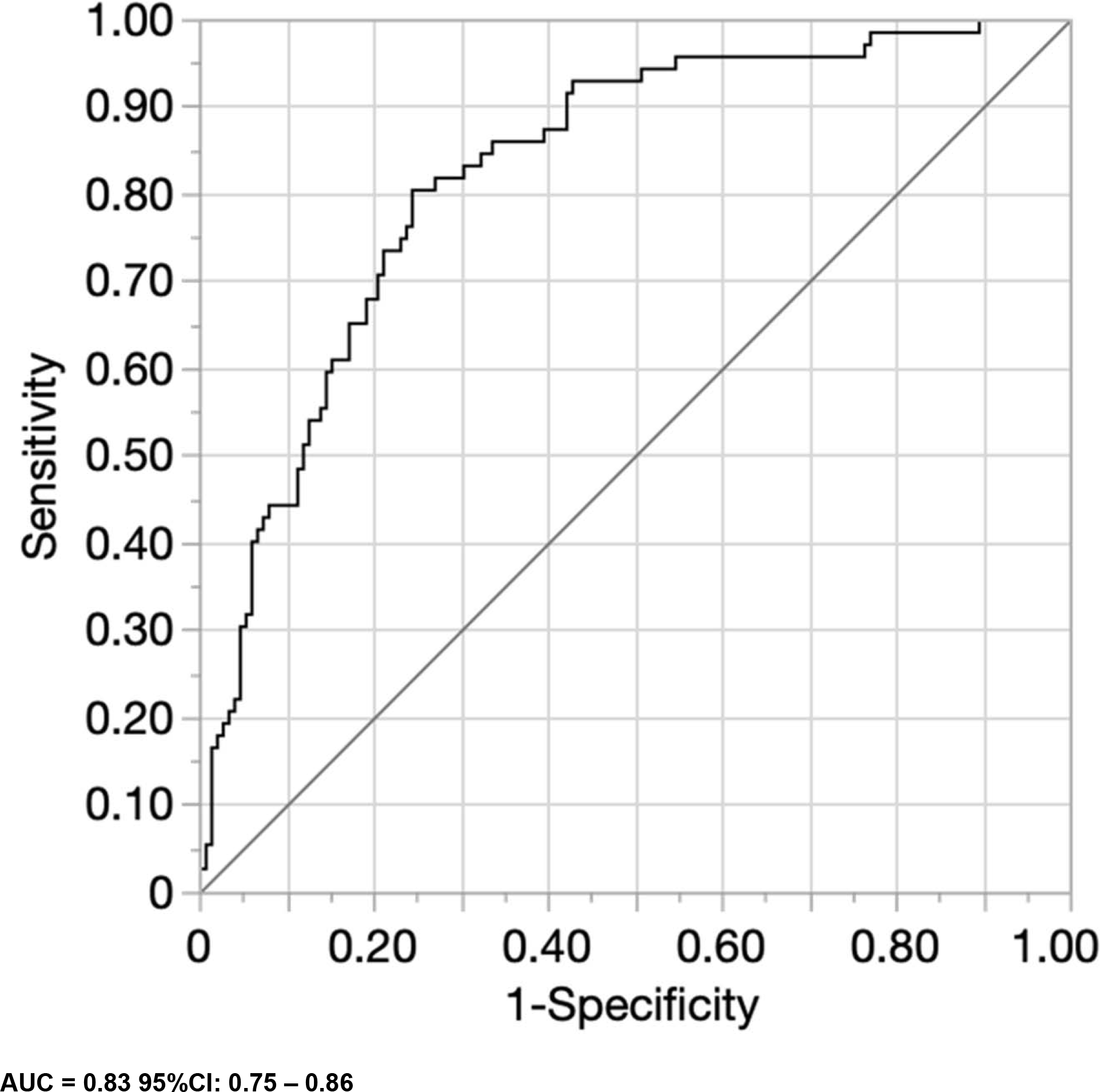
Receiver operator curve analysis of the combined multivariate logistic regression model with age, admission NIHSS, rCBF < 30% volume, mTICI score, admission glucose, and sex.

## Discussion

Our study demonstrates that excellent recanalization, smaller core volumes, lower age, and decreased initial stroke severity are all independent predictors of favorable outcomes as measured by AMPAC. Our combined model with the aforementioned factors demonstrated strong performance (AUC 0.83 [95% CI 0.75 - 0.86]). Our results support the emphasis on minimizing ischemic core volume, while also underscoring the importance of maximizing chances of excellent recanalization in order to optimize functional abilities of patients with AIS-LVO. To our knowledge, this the first study to investigate the potential of pretreatment imaging parameters in predicting favorable AMPAC scores.

Standard of care guidelines have prioritized the identification of optimal post-acute care for stroke patients^8,19,20^. Early discharge planning is essential for AIS-LVO patients in order to optimally utilize the finite resources within the hospital and subsequent rehabilitation settings^8^. Furthermore, predicting whether patients can be discharged home instead of a post-acute care setting (acute rehabilitation, subacute, rehabilitation, etc.) has important ramifications for the patients’ psychosocial profile including cognition, stroke recurrence prevention, insurance status, availability, and treatment of comorbid conditions^4,5,8,19^. This complex and nuanced decision-making process can therefore be aided by pretreatment predictors of discharge status for early planning in AIS-LVO patients.

Our study suggests that pretreatment comprehensive CT imaging may also be useful adjunct tools in this challenging discharge planning process. Although ischemic core volume in AIS-LVO as a predictive biomarker is well established with widely used functional outcome measures such as modified Rankin score (mRS)^13,14,21^, determining the relationship between ischemic core with AMPAC was yet to be performed. Our findings demonstrate that smaller core volumes based on rCBF < 30% volumes independently predict favorable AMPAC scores. Although the association of smaller ischemic core volumes with better outcomes is an expected finding, it also adds significance to the importance of smaller cores, as this result also affects functional ability and early discharge planning.

Another significant parameter predictive of favorable AMPAC scores is achieving excellent recanalization by MT (defined as mTICI 2c/3). Several landmark trials in 2015 in the early window^22–25^ and later in 2018 for the late window^14,21^ established MT as the standard of care for AIS-LVO. Additional studies have also demonstrated that achieving excellent recanalization^26–30^ further improves outcomes compared to mTICI 2b, despite also being considered successful recanalization. We found a higher likelihood of achieving favorable AMPAC scores with excellent recanalization (mTICI 2c/3) compared to mTICI 2b or lower (OR 3.06). Our study is concordant with prior trials and subsequent studies, corroborating the efficacy of MT and the need to achieve excellent recanalization to maximize likelihood of favorable functional outcomes. Our work extends these established outcome related findings by emphasizing the importance of excellent recanalization with discharge planning as well.

Analyses of other baseline characteristics also confirmed some expected findings. Younger patients and patients who presented less severely with AIS were also associated with favorable functional outcomes as measured using AMPAC. Both of these factors are well known predictors of improved outcomes in AIS-LVO patients. Younger AIS-LVO patients, especially those under 50, tend to have fewer postprocedural complications and better outcomes compared to patients 50 years or older^31^. Lower initial stroke severity, similarly, is a well-established independent predictor of better outcomes in AIS-LVO^31^. Our results are therefore concordant with prior studies identifying these factors as long-standing biomarkers of clinical outcomes.

We acknowledge some limitations in this study. First, this study is inherently limited by its retrospective approach. Secondly, CTP is not widely available in smaller and rural centers, thus limiting generalizability. Prospective studies are needed to validate these findings.

## Conclusion

Smaller core volumes, younger age, lower initial stroke severity, and excellent recanalization are significantly predictive of favorable functional outcomes as measured using AMPAC. Our study further emphasizes the significance of minimizing core volume and aiming for excellent recanalization in order to optimize functional outcomes for discharge planning in AIS-LVO patients.

## Data Availability

Data can be made available upon reasonable request

## Notes

### Competing Interest Statement

Drs. Greg Albers, Jeremy Heit, and Vivek Yedavalli are consultants for Rapid (iSchemaView, Menlo Park, CA)

### Clinical Trial

N/A

### Funding Statement

No funding to disclosure

### Author Declarations

This study was approved through the Johns Hopkins School of Medicine institutional review board (JHU-IRB00269637).

